# Evaluation of the Nallasamy Formula: A Stacking Ensemble Machine Learning Method for Refraction Prediction in Cataract Surgery

**DOI:** 10.1101/2021.10.25.21265489

**Authors:** Tingyang Li, Joshua D. Stein, Nambi Nallasamy

**Affiliations:** Department of Computational Medicine and Bioinformatics, University of Michigan; Kellogg Eye Center, Department of Ophthalmology and Visual Sciences, University of Michigan; Center for Eye Policy and Innovation, University of Michigan, Ann Arbor, MI; Department of Health Management and Policy, University of Michigan School of Public Health, Ann Arbor, MI

## Abstract

**Aims:** To develop a new intraocular lens (IOL) power selection method with improved accuracy for general cataract patients receiving Alcon SN60WF lenses.

**Methods and Analysis:** A total of 5016 patients (6893 eyes) who underwent cataract surgery at University of Michigan’ s Kellogg Eye Center and received the Alcon SN60WF lens were included in the study. A machine learning-based method was developed using a training dataset of 4013 patients (5890 eyes), and evaluated on a testing dataset of 1003 patients (1003 eyes). Each eye had a complete profile of preoperative biometry, the implanted IOL power, and postoperative refraction. The performance of our method was compared to that of Barrett Universal II, Haigis, Hoffer Q, Holladay 1, and SRK/T.

**Results:** MAE of the Nallasamy formula in the testing dataset was 0.312 Diopters (MedAE = 0.242 D). Performance of existing methods were as follows: Barrett Universal II MAE = 0.328 D (MedAE = 0.256 D), Haigis MAE = 0.363 D (MedAE = 0.289 D), Hoffer Q MAE = 0.404 D (MedAE = 0.331 D), Holladay 1 MAE = 0.371 D (MedAE = 0.298 D) and SRK/T MAE = 0.376 D (MedAE = 0.300 D). The Nallasamy formula performed significantly better than all five existing methods based on the paired Wilcoxon test with Bonferroni correction (p-value < 0.05).

**Conclusions:** Nallasamy formula outperformed the five methods studied (including Barrett Universal II) on overall MAE and MedAE, percentage of eyes within 0.5 D of prediction, as well as MAE in short, medium, and long axial length eyes.

**SYNOPSIS:** Nallasamy formula, a novel machine learning-based IOL power calculation formula developed based on a dataset of 6893 eyes, achieved significantly better prediction accuracy than five traditional IOL power formulas including Barrett Universal II.

## INTRODUCTION

Cataract surgery is the most commonly performed surgical procedure in the United States (approximately 4 million/year) and worldwide (approximately 23 million/year). The appropriate selection of IOL power based on accurate prediction of postoperative refraction is necessary for achieving a favorable refractive outcome and is closely associated with patient satisfaction. An inappropriate IOL power was found to be the indication for approximately 20% of cataract surgery cases that required secondary intervention, lens removal or lens exchange, according to analyses of records between 2002 and 2017.[1,2]

Various generations of IOL power calculation formulas have been published since the 1960s. From the earliest regression formulas (Binkhorst formula, SRK formula) to the fourth and fifth generation of vergence formulas which established the effective lens position (ELP) as a function of the axial length, lens thickness and keratometry, the accuracy of IOL power calculation has been substantially improved. Among all conventional formulas, the Barrett Universal II formula[3] is considered to be the current gold standard.

Although the methodology for IOL power selection has been studied for decades, patient expectations for refractive outcomes continue to rise and room remains for improvement in refraction prediction performance. Machine learning (ML) and artificial intelligence have proven to be successful in many medical applications, including ophthalmology.[4,5] Researchers have begun to incorporate ML into IOL power calculations in recent years.

However, key limitations exist among recently-published ML-based IOL calculation methods: (1) performance comparisons limited to older generation formulas,[6] (2) failure to achieve statistically significant improvement over current generation formulas,[7] and (3) small datasets that leave the robustness and generalizability of methods in question.[8]

With a goal of advancing the understanding of IOL power selection for general cataract patients and improving refraction prediction accuracy, in the presented study, we developed a novel machine learning-based IOL power calculation method, the Nallasamy formula, based on a large dataset of 5016 cataract patients. In this model, we employed ensemble machine learning methods and novel data augmentation methods. The method reported here performed statistically significantly better than the formulas studied, including Barrett Universal II, on an unseen testing dataset of 1003 patients.

## MATERIALS AND METHODS

### Data collection and preprocessing

The presented study focused on a subset of patients receiving care at the University of Michigan between August 25, 2015 and June 27, 2019. The preoperative biometry records were obtained from Lenstar LS 900 optical biometers (Haag-Streit USA Inc, EyeSuite software version i9.1.0.0) at University of Michigan’ s Kellogg Eye Center. Patient demographics (including patient age, gender, and ethnicity) and cataract surgery information were obtained via the Sight Outcomes Research Collaborative (SOURCE) Ophthalmology Data Repository. SOURCE is a data repository that tracks the electronic health record (EHR) data of all patients receiving any eye care at participating academic medical institutions. The information deposited in SOURCE includes patient demographics, diagnoses identified based on International Classification of Diseases (ICD) codes, procedures based on Current Procedural Terminology (CPT) codes, and structured and unstructured (free-text) data from all clinical encounters (clinic visits, operative reports, etc.). Various studies using data from SOURCE were published.[9–13] Institutional review board approval was obtained for the presented study. All subjects were fully anonymized and, therefore an informed consent was not required for this retrospective study. The study was carried out in accordance with the tenets of the Declaration of Helsinki.

The inclusion criteria for the cases were as follows: (1) Cataract surgery was performed (CPT code = 66982 or 66984). (2) An Alcon SN60WF one-piece acrylic monofocal lens was implanted, (3) No refractive surgery was performed before the cataract surgery. (4) No additional surgery was performed at the time of cataract surgery. Cases with any CPT code other than 66982 or 66984 were excluded. (5) Visual acuity was 20/40 or better. (6) Data was complete and was not out of bounds for any of the five formulas with which performance was compared.

### Stacking ensemble machine learning framework

After all preprocessing steps, we obtained a clean tabular dataset of 5016 patients wherein each eye had a complete profile of preoperative biometry, patient demographics (patient gender and age), the power of the implanted IOL and the postoperative refraction. Preoperative biometry included the axial length (AL), crystalline lens thickness (LT), anterior chamber depth (ACD), aqueous depth (AD), astigmatism, white-to-white (WTW), central corneal thickness (CCT), and keratometry (K1 and K2, *K* = (*K*1 + *K*2)/2). The postoperative refraction was calculated from the spherical component (SC) and the cylindrical component (CC) with an adjustment with regard to the lane length at Kellogg Eye Center (10 feet, 3.048 meters): spherical equivalent (SE) *refraction* = (SC − 0.1614) + 0.5CC according to Simpson and Charman’ s recommendation.[14]

The prediction task was framed as a regression problem where the goal was to build a machine learning algorithm that predicts the postoperative refraction using available information. The value to be predicted is referred to as the target value (represented as Y in **Figure 1**) and the inputs that are used to make the predictions are referred to as features or predictors (represented as X in **Figure 1**). The dataset was randomly split into a training/validation set with 4013 patients (5890 eyes) which was 80% of all patients, and a testing set with 1003 patients which was 20% of all patients (**Figure 1**). In order to make sure all samples in the testing set were independent, one eye was selected at random and dropped from the dataset for all patients with both eyes available in the dataset. The training/validation set was used for cross-validation and hyperparameter selection of the machine learning model. The testing set was used for performance comparison between the existing conventional formulas and our ML-based method.

**Figure 1.**
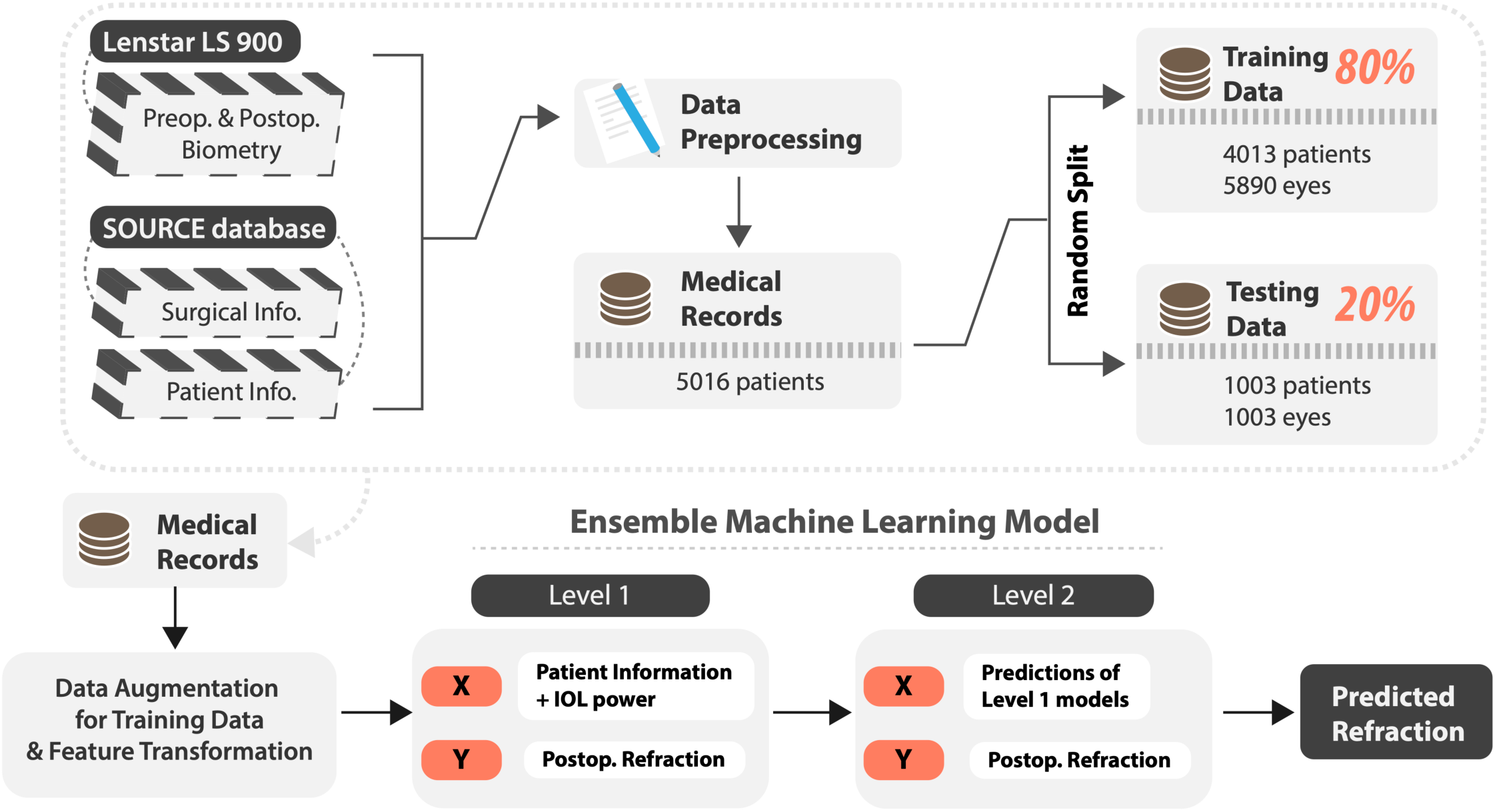
The overall method pipeline.

Ensemble learning is a technique that involves combining the predictions from base learners with the goal of reducing variance and achieving improved prediction performance. An ensemble model is usually believed to outperform individual learners in most cases.[15] Stacking (or stacked generalization) is one of the most commonly used meta-learning paradigms, where a number of base-learners are trained using the raw training data and a single meta-learner is trained to combine the predictions from the base-learner.[16] The reason for using an ensemble ML model in this study is to take advantage of different classes of ML algorithms and improve the overall performance of the model. The stacking model consists of two layers. In the first layer a group of level-1 learners was trained based on the raw data (preoperative patient data and the postoperative refraction). The second layer consists of the metamodel which uses the output of the level-1 learners as the input features. Therefore, the number of input features for the level-2 model equals the number of level-1 models. The output from the level-2 meta-model is the final prediction result (**Figure 1**).

### Lens constant optimization of conventional methods

The conventional formulas Haigis, Hoffer Q, Holladay 1, SRK/T were implemented in Python based on their specific equations.[17–24] The prediction results of Barrett Universal II were obtained through the online calculator.[3] The constants of the corresponding formulas were optimized based on the cases in the training dataset (4013 patients). The most optimal constant was selected by zeroing the mean prediction error. The optimized constants are listed in **Table 1**.

**Table 1.**
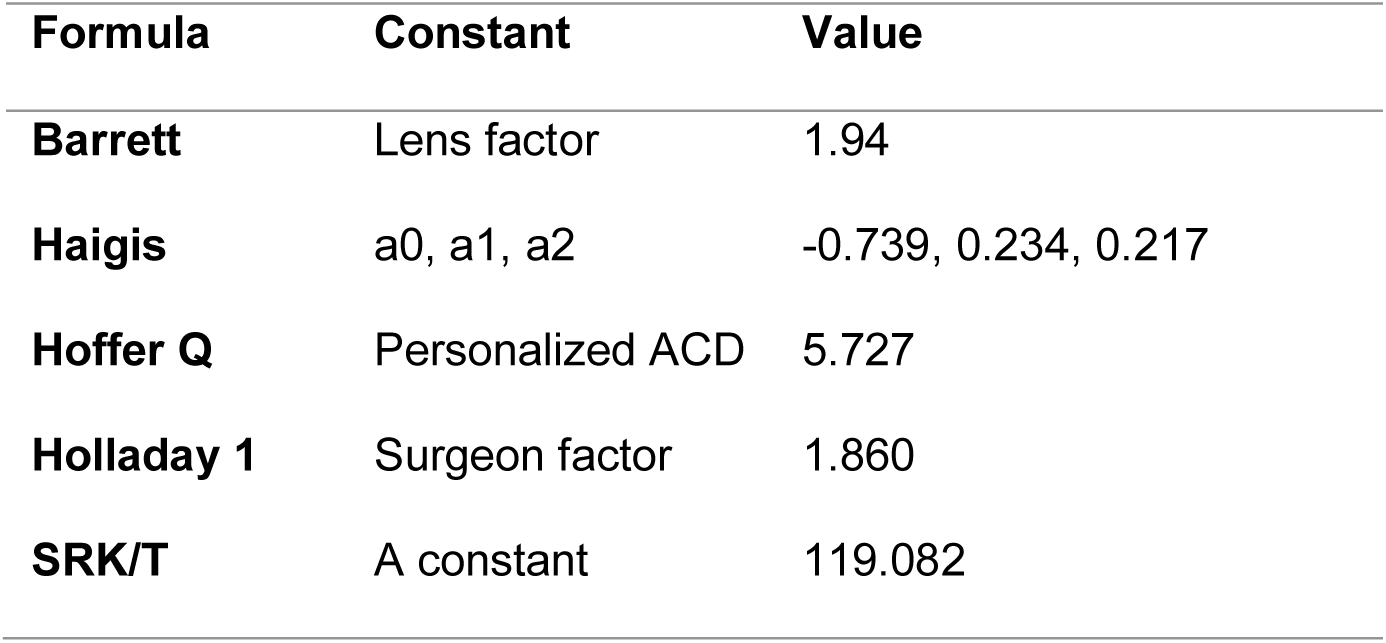
The optimized lens constants.

### Cross-validation and hyperparameter tuning

During the development of the ML model, we performed model evaluation and selection through five-fold cross-validation. During the cross-validation, 4013 training/validation cases were divided into training sets and validation datasets. A random eye was removed for patients with both eyes available in the validation dataset. The optimization of hyperparameters of the machine learning models, the combination of the level-1 models, and the selection of the level-2 model were performed by minimizing the averaged mean absolute error (MAE) based on the cross-validation results.

### Performance comparison on the testing set

To compare the performance between our method and conventional IOL formulas, we trained the ML-model with the entire training dataset (5890 eyes) and made predictions on the testing dataset. We calculated the mean arithmetic error (ME), mean absolute error (MAE), median absolute error (MedAE) of the postoperative refraction predictions and the standard deviation (SD) of the prediction error. We also calculated the number and percentage of patients with an absolute prediction error of less than or equal to 0.25 D, 0.50 D, 0.75 D, and 1.00 D, and evaluated the statistical significance of the difference between formulas with Cochran’ s Q test. The statistical significance of the difference between the testing set performance of the IOL formulas was assessed using a Friedman test followed by a paired Wilcoxon test with Bonferroni correction. To investigate the performance of our method in cases with different axial lengths, we calculated the SD, ME, MAE, and MedAE for patients in the short AL group (AL < 22 mm), medium AL group (22 mm ≤ AL ≤ 26 mm) and long AL group (AL > 26 mm). In addition to the above metrics, we calculated the slope of the correlation between the arithmetic error and AL as *m*. Using the above variables, we computed the IOL Formula Performance Index (FPI) as recommended by Hoffer et al[25] for each formula as follows, where *n* is the percentage of eyes with an absolute error within 0.5 D. Higher FPI means better accuracy.

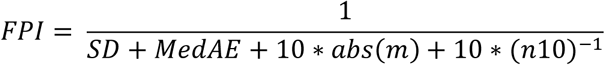

To investigate the effect of the size of the training data on the performance of the ML model, we randomly sampled 10%, 20%, …, 90% of the training data, then retrained and compared the alternative models’ results on the testing set. The proportions of training cases were adjusted before the application of data augmentation and data transformation techniques. All other configurations and hyperparameters were kept the same for alternative models except for the number of training cases.

In this study, the refraction prediction error was defined as follows. The criterion for statistical significance was p-value < 0.05. All statistical analyses were scripted with Python 3.9.5.

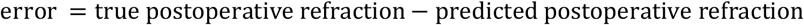

## RESULTS

Out of 5016 patients, 4013 patients (5890 eyes) were assigned to the training/validation dataset, and 1003 cases were isolated as a hold-out testing dataset for performance comparison. A summary of the patient demographics in the training and testing sets is shown in **Table 2**. The distribution of data is shown in **Figure S1**.

**Table 2.** Summary of patient demographics. SD: standard deviation; K: keratometry; AL: axial length; LT: lens thickness; ACD: anterior chamber depth; D: diopter.

| Characteristic | Training set (mean ± SD) | Testing set (mean ± SD) |
| --- | --- | --- |
| Count | 5890 eyes, 4013 patients | 1003 eyes, 1003 patients |
| Gender | Male: 2573 eyes (43.7%),<br>Female: 3317 eyes (56.3%) | Male: 433 eyes (43.2%),<br>Female: 570 eyes (56.8%) |
| Age at surgery (years) | 71.00 ± 9.43 | 70.73 ± 9.50 |
| Preoperative K (D) | 43.87 ± 1.54 | 43.88 ± 1.56 |
| Preoperative AL (mm) | 24.17 ± 1.34 | 24.15 ± 1.35 |
| Preoperative LT (mm) | 4.53 ± 0.44 | 4.53 ± 0.45 |
| Preoperative ACD (mm) | 3.26 ± 0.41 | 3.25 ± 0.41 |
| Postoperative refraction (D) | -0.55 ± 0.85 | -0.59 ± 0.93 |

The performance of our method and conventional methods is shown in **Table 3**. According to the Wilcoxon test, our method performed significantly better than all the other five methods with an MAE of 0.312 D, which was 4.9% lower than that of Barrett (0.328 D). The specific p-values can be found in **Table S1**. Our method also achieved the highest Formula Performance Index (FPI).

**Table 3.** Performance summary in the testing set. MAE: mean absolute error; MedAE: median absolute error; ME: mean error, SD: standard deviation of the prediction error; m: the axial length bias, computed as the slope of the correlation between the arithmetic error and AL; AE ≤ 0.5 D: percentage of eyes with an absolute error (AE) less than or equal to 0.5 D; FPI: Formula Performance Index. The unit for the errors is Diopter (D). Wilcoxon test p-value < 0.05 indicates the statistical significance of the difference between the performance of our method and a conventional method.

| | MAE | MedAE | ME | SD | m | AE $\leq 0.5$ D | FPI | P-value |
| --- | --- | --- | --- | --- | --- | --- | --- | --- |
| **Barrett** | 0.328 | 0.256 | 0.038 | 0.437 | 0.307 | 78.3% | 0.198 | p < 0.05 |
| **Haigis** | 0.363 | 0.289 | 0.024 | 0.469 | 0.226 | 74.7% | 0.230 | p < 0.05 |
| **Hoffer Q** | 0.404 | 0.331 | 0.009 | 0.518 | 0.951 | 70.3% | 0.085 | p < 0.05 |
| **Holladay 1** | 0.371 | 0.298 | 0.021 | 0.487 | 0.773 | 74.0% | 0.101 | p < 0.05 |
| **SRK/T** | 0.376 | 0.300 | 0.014 | 0.485 | 0.486 | 73.2% | 0.143 | p < 0.05 |
| **Our Method** | 0.312 | 0.242 | 0.015 | 0.418 | -0.033 | 80.2% | 0.447 | / |

The percentage of patients with an absolute error less than or equal to 0.25 D, 0.50 D, 0.75 D, and 1.00 D is shown in **Figure 2**. Our method resulted in a larger percentage of patients in the absolute error ≤ 0.5 D group (80.2%) compared to Barrett (78.3%), and a larger percentage of patients in the absolute error ≤ 1.0 D group (97.6%) compared to Barrett (96.6%). Overall, our method achieved the highest percentage in the absolute error ≤ 0.5 D group among all six formulas, and was statistically better than Barrett (Cochran’ s Q test p-values were shown in **Table S2**).

**Figure 2.**
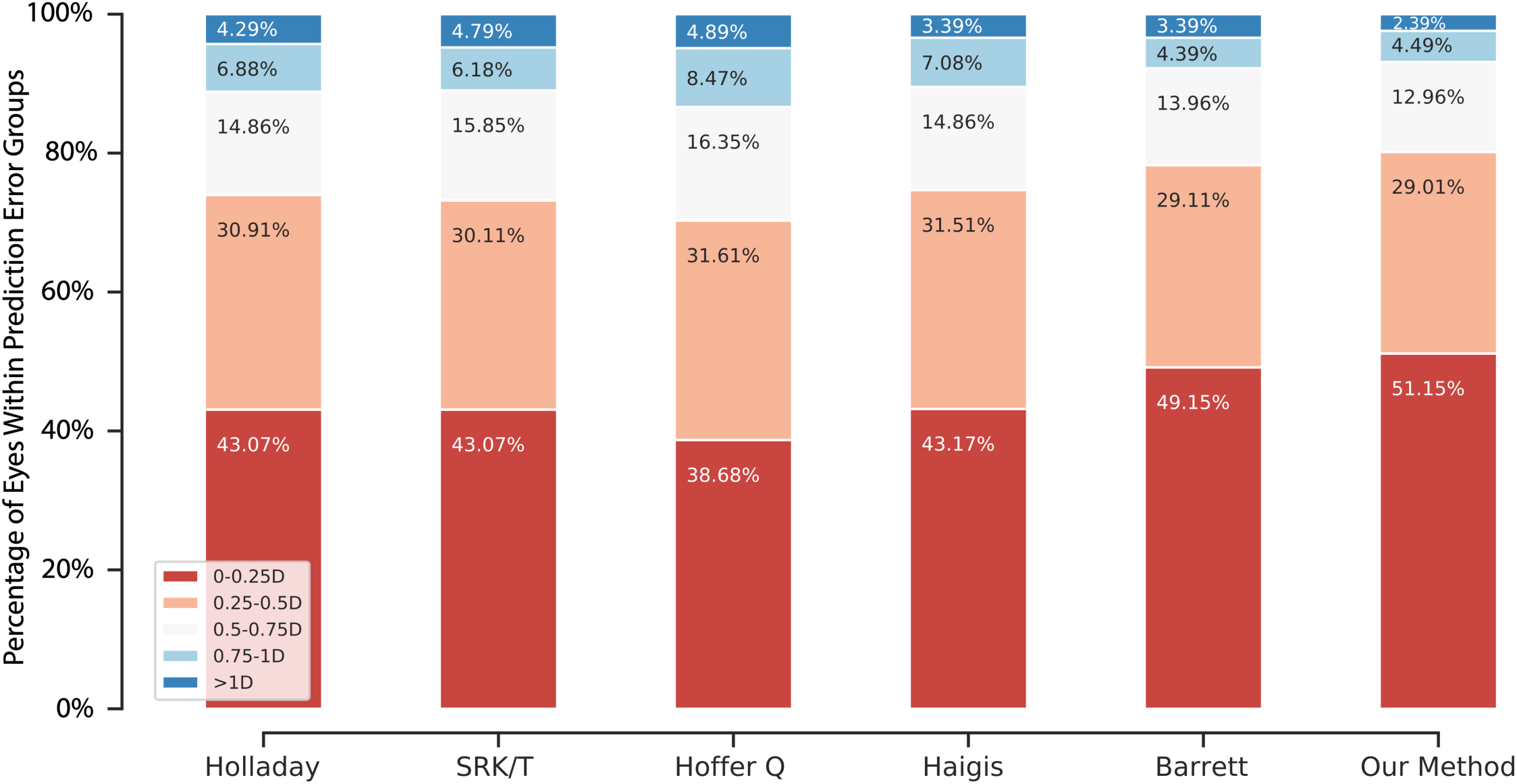
The percentage of patients in each error category for each formula, calculated based on the results in the testing dataset.

We compared the performance of the tested formulas among patients with different axial lengths in **Table 4**. Numerically, our method achieved the lowest MAEs and SDs among all 6 formulas in all 3 AL groups. The relationship between the prediction errors and the ALs is shown in **Figure 3**. The errors of our method remained close to zero across the whole span of ALs.

**Table 4.**
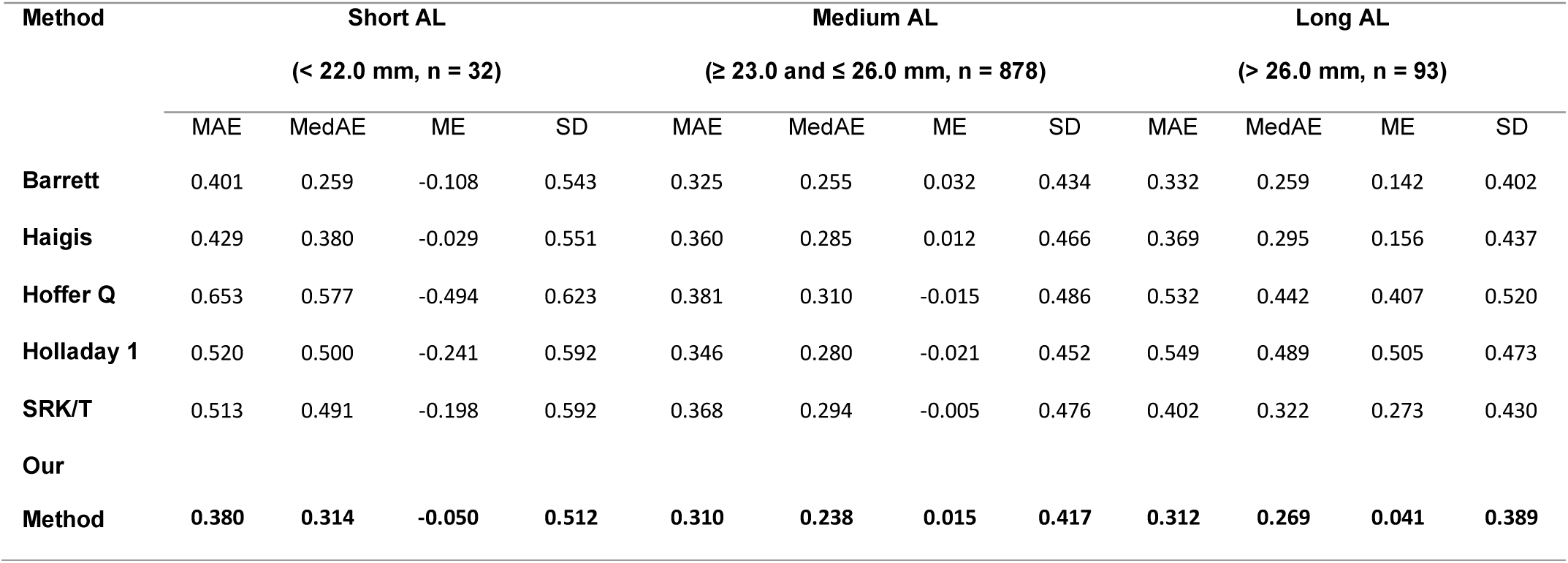
The postoperative refraction prediction performance of five existing formulas and our method in short/medium/long AL groups in the testing set. AL: axial length; n: number of eyes in each group; MAE: mean absolute error; MedAE: median absolute error; ME: mean error; SD: standard deviation of the prediction error.

**Figure 3.**
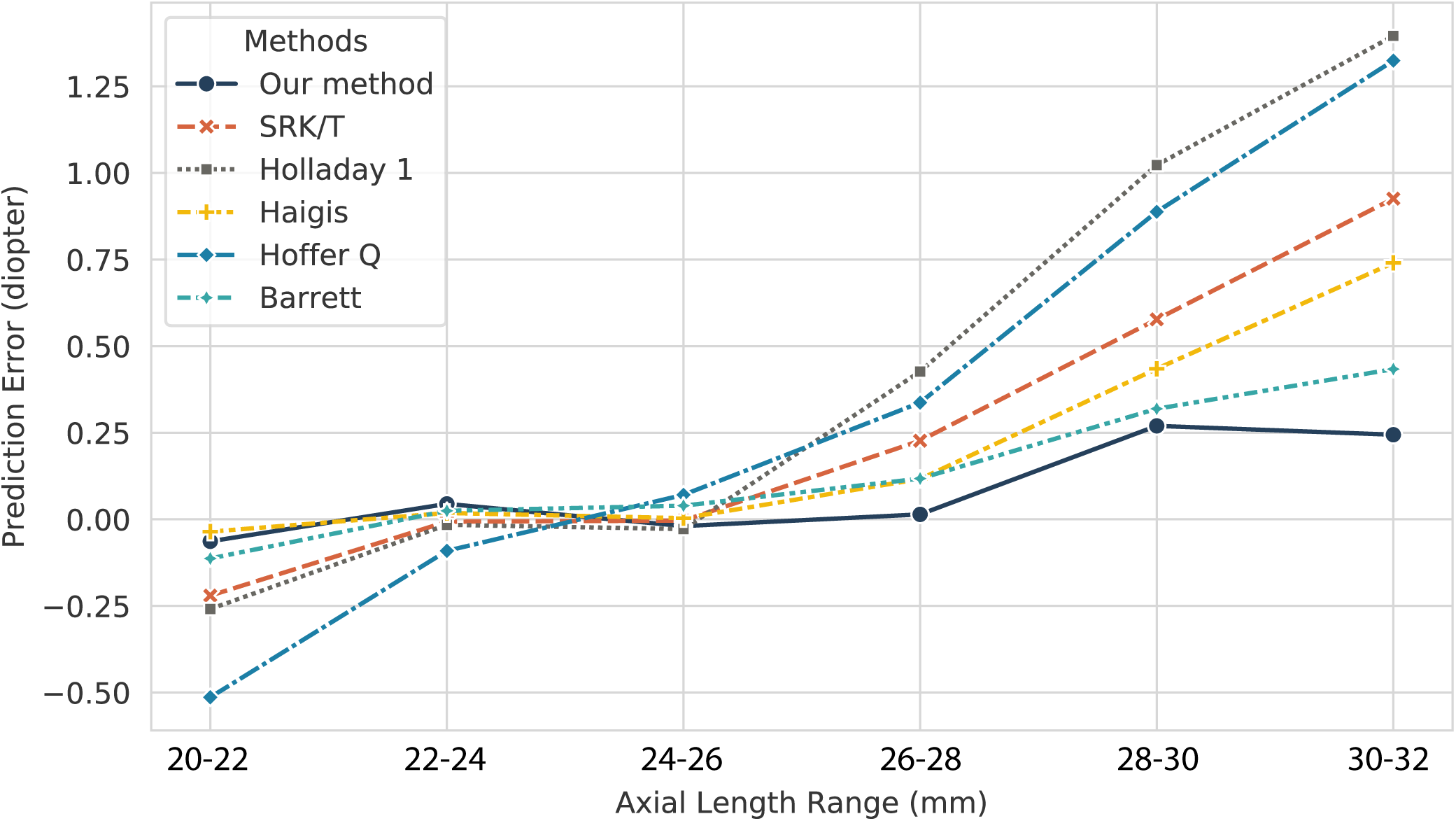
The mean prediction errors in the testing set grouped based on axial lengths. Each dot represents the mean prediction error of eyes with an axial length between a specific range.

When the model was trained with different proportions of the training data (**Figure 4**), the corresponding performance on the testing set displayed a trend toward improving performance (decreased MAE) with increasing training set sizes.

**Figure 4.**
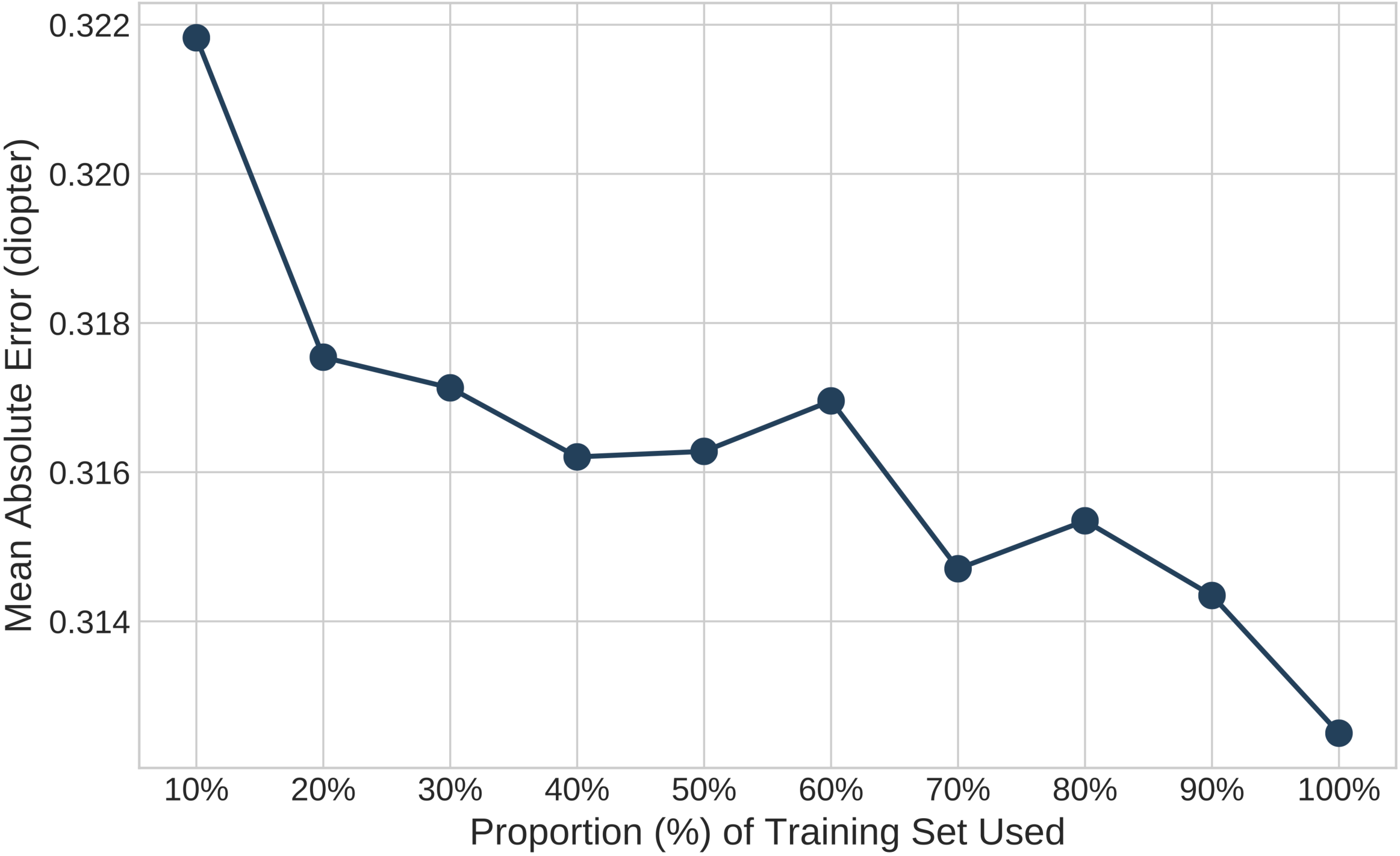
The change of the mean absolute prediction error in the testing set when the machine learning method uses 10%, 20%, …, 100% of the training data.

## DISCUSSION

We have presented here a new machine learning-based IOL power calculation method which performs statistically significantly better than Barrett Universal II on a large unseen testing dataset. We chose an ensemble machine learning framework for this particular problem, and this choice allows the method to compensate for the potential biases of individual learners. During the development of the model, we designed and applied several data augmentation methods to enhance prediction performance. Data augmentation methods are not only beneficial for enlarging the dataset size, but also to address natural imbalances in clinical datasets. The biometry measures are not uniformly distributed as shown in **Figure S1**. For example, the axial length has more instances in the medium group (between 22 mm and 26 mm) compared to the long and short AL groups. The postoperative refractions and the implanted IOL powers were not uniformly distributed either. All IOL powers in the dataset were manually selected by surgeons with a particular target refraction in mind, typically between 0 D and -3 D. Data augmentation helps to account for the scarcity of extreme cases and biases introduced by clinical decision-making process.

In this study, we used a relatively large dataset of 6893 eyes. Evaluation of the relationship between the proportion of the available training data used and MAE demonstrated the expected inverse relationship. This trend continued even as the training set was increased from 90% to 100% of the available training data (**Figure 3**), indicating the potential for further improvement as the same model is exposed to larger datasets.

We achieved lower MAEs than Barrett Universal II in all three axial length groups. Our method yielded 80.2% of eyes with a predicted refraction within 0.5 D of the true refraction, which was approximately 2% more than that of Barrett (78.3%). This difference was statistically significant and, considering the 23 million cataract surgeries performed per year worldwide, is likely to be of clinical significance.

Recently, Hoffer et al. proposed in *Ophthalmology* the use of the Formula Performance Index (FPI) as a means of evaluating and ranking the performance of IOL power calculation methods.[25] Higher values of the FPI indicate higher performance. Our method strongly outperformed the conventional formulas on FPI, achieving a 0.447 FPI while the conventional formulas ranged from 0.085 to 0.230 (**Table 3**). The FPI takes into account the (1) SD of the prediction error, (2) the MedAE, (3) the AL bias, and (4) the percentage of eyes with refraction predictions within 0.5 D of true refractions. Our method demonstrated superior performance on each of these individual metrics, as summarized in **Table 3**. Of particular note is our method’ s superior SD of the prediction error, which Holladay et al. recently referred to as “the single best parameter to characterize the performance of an IOL power calculation formula.”[26]

Also of interest is the AL bias, which is calculated as the slope of the correlation of the AL and the prediction error for a given formula. Conventional IOL formulas demonstrate strong correlations between AL and the prediction error, as depicted in **Figure 3**. Machine learning-based methods such as ours, on the other hand, have the potential to better capture the nonlinearity of the relationship between biometric variables, IOL power, and postoperative refraction, resulting in substantially smaller AL bias (e.g., -0.03 for Nallasamy vs. 0.31 for Barrett). This translates to improved performance across AL categories (short, medium, and long), and should obviate the need for using different formulas based on axial length.

We are aware of multiple limitations of our study. Our method has not yet been validated on a dataset from a different medical institution. Performance analysis on external datasets will be a focus of future work as we begin to apply our approach to different populations around the world. Another limitation is that we were not able to compare our performance with a few formulas such as Hill-RBF because of a lack of access. However, prior studies indicate that Barrett Universal II is a good reference point for top-tier IOL formulas.[27–29] An additional limitation is that at present, our method has been customized for the Alcon SN60WF lens, and additional data will be needed to adjust the method for additional lens models.

An intrinsic difference between ML-based methods and the vergence formulas is that vergence formulas estimate the effective lens position (ELP) as a vital variable during the calculation of the postoperative refraction, but ML-based methods usually take a one-step approach for prediction, unless the model is specifically designed to predict both the ELP and the postoperative refraction. In previously published work, we reported the development of an ML-based method for postoperative anterior chamber depth (ACD) estimation.[11,12] However, the method presented here does not rely upon prediction of a postoperative ACD or ELP as an intermediate variable, unlike the vergence formulas. This approach may allow the ML method to avoid the propagation of errors (however small) introduced during the prediction of the postoperative ACD or ELP.

While the theoretical optics-based methods remain crucial for special cases, machine learning offers improved performance for large populations through the identification of latent patterns in historical data that can go unrecognized by conventional methods. To that end, we have reported here the successful development and testing of a machine learning-based approach to IOL power calculation for cataract surgery that outperforms Barrett Universal II on all broadly accepted metrics of IOL calculation performance. The Nallasamy formula will be made freely available to the public to use online.

## Data Availability

Data are not publically available.

## ACKNOWLEDGMENT

None

## CONTRIBUTIONS

TL: data analysis, programming, and writing of the manuscript; JDS: data collection; NN: data analysis, programming, data collection, guidance on method development, and writing of the manuscript

## FUNDING

This work was supported by the Lighthouse Guild, New York, NY (JDS) and National Eye Institute, Bethesda, MD, 1R01EY026641-01A1 (JDS).

## COMPETING INTERESTS

None declared

## DATA AVAILABILITY STATEMENT

Data are not publicly available

## PATIENT AND PUBLIC INVOLVEMENT

It was not appropriate or possible to involve patients or the public in the design, or conduct, or reporting, or dissemination plans of our research.

## Notes

### Competing Interest Statement

The authors have declared no competing interest.

### Funding Statement

This study was funded by the Lighthouse Guild, New York, NY (JDS) and National Eye Institute, Bethesda, MD, 1R01EY026641-01A1 (JDS).

### Author Declarations

IRB of the University of Michigan gave ethical approval for this work.

