## Supplementary material for "Evaluation of the Nallasamy Formula: A Stacking Ensemble Machine Learning Method for Refraction Prediction in Cataract Surgery": Figure S1

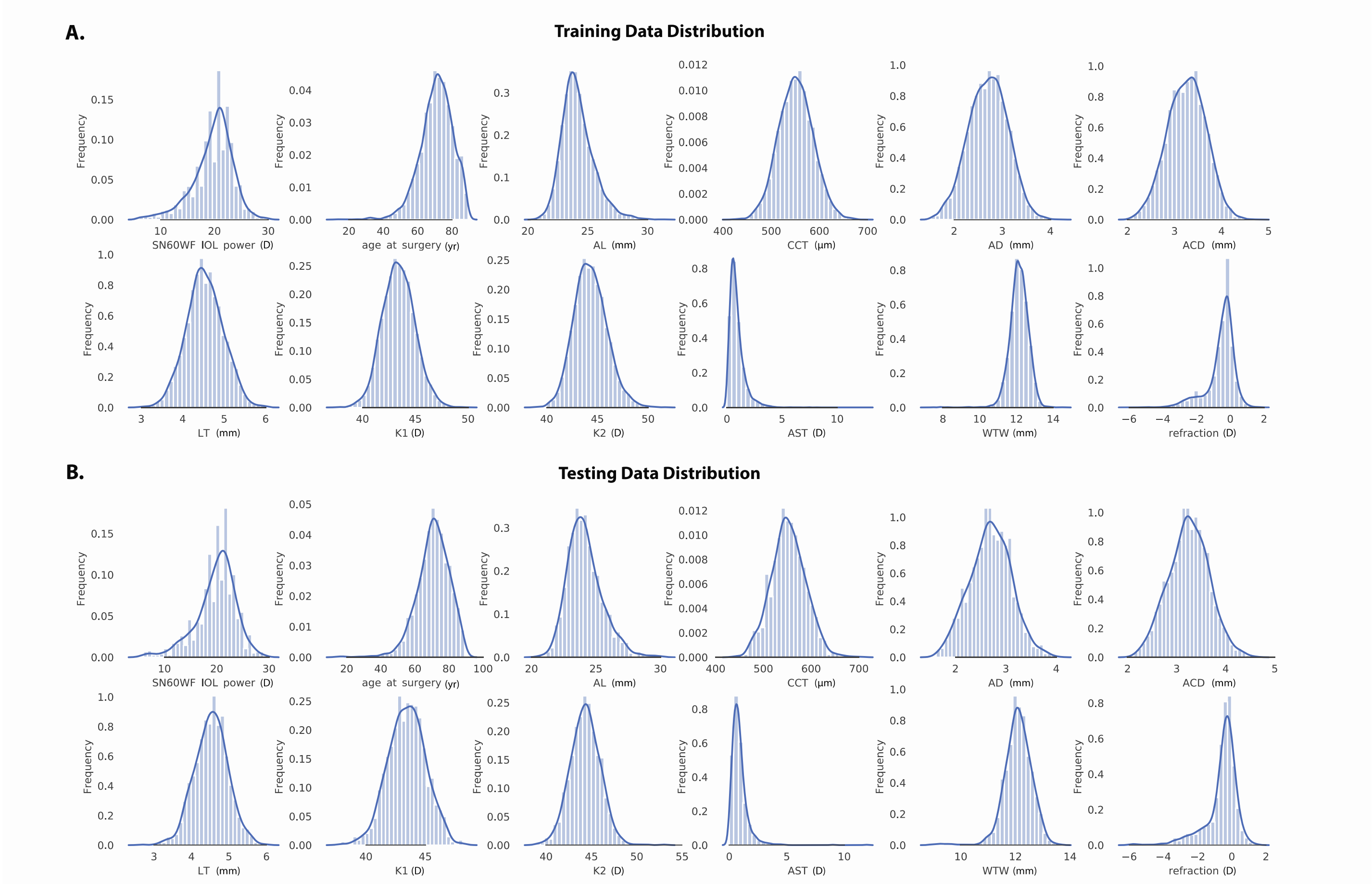


**Figure S1** Distribution of data in the training and testing datasets. **A.** Training data distribution. **B.** Testing data distribution. The number of bins for the bar plots is 30. The curve in each plot is a gaussian kernel density estimate of the distribution. AL: axial length; CCT: central corneal thickness; AD: aqueous depth; ACD: anterior chamber depth; LT: lens thickness; K1: flat keratometry; K2: steep keratometry, AST: astigmatism; WTW: white-to-white; D: diopter
