## Supplementary material for "Evaluation of the Nallasamy Formula: A Stacking Ensemble Machine Learning Method for Refraction Prediction in Cataract Surgery": Table S1

| Formula | Our Method | SRK/T | Holladay 1 | Haigis | Hoffer Q |
|---|---|---|---|---|---|
| **SRK/T** | 1.45e-18 | / | / | / | / |
| **Holladay 1** | 4.99e-14 | 6.82e-01 | / | / | / |
| **Haigis** | 5.10e-15 | 1.00e+00 | 1.00e+00 | / | / |
| **Hoffer Q** | 6.74e-24 | 4.15e-02 | 6.65e-09 | 1.39e-09 | / |
| **Barrett** | 5.76e-04 | 3.76e-14 | 1.61e-09 | 5.92e-09 | 1.06e-22 |

**Table S1** The p-values from the post-hoc paired Wilcoxon tests, following the Friedman test, for the comparison of the testing set performance between methods. The p-values were adjusted with Bonferroni correction. The p-values between our method and the conventional methods were shown in the first column. All significant p-values (p-value < 0.05) were underscored. The Friedman test statistic was 176.73, and the associated p-value was 2.67e-36, which was statistically significant.
