## Supplementary material for "Evaluation of the Nallasamy Formula: A Stacking Ensemble Machine Learning Method for Refraction Prediction in Cataract Surgery": Table S2

| Formula | Within 0.25 D | Within 0.50 D | Within 0.75 D | Within 1.00 D |
|---|---|---|---|---|
| **Vs. Barrett** | 1.18e-1 | 4.16e-2 | 8.33e-2 | 1.24e-2 |
| **Vs. Haigis** | 2.06e-06 | 9.98e-06 | 4.83e-06 | 1.84e-2 |
| **Vs. Hoffer Q** | 3.70e-11 | 3.37e-11 | 2.25e-10 | 3.96e-05 |
| **Vs. Holladay 1** | 4.83e-07 | 8.12e-07 | 9.63e-08 | 2.56e-4 |
| **Vs. SRK/T** | 1.89e-06 | 9.43e-08 | 7.98e-07 | 5.74e-06 |

**Table S2** The p-values from Cochran's Q test for the comparison of number cases within 0.25 D, 0.50 D, 0.75 D, and 1.00 D between our method and conventional methods in the testing set. All significant p-values (p-value < 0.05) were underscored. D: diopter.
